# Common and Disorders-Specific Cortical Thickness Alterations in Internalizing, Externalizing and Thought Disorders over a 2-year Period in the Preadolescents of the ABCD Study

**DOI:** 10.1101/2021.09.02.21263005

**Authors:** Gechang Yu, Xinran Wu, Zhaowen Liu, Benjamin Becker, Kai Zhang, Nanyu Kuang, Jujiao Kang, Guiying Dong, Xing-Ming Zhao, Gunter Schumann, Jianfeng Feng, Barbara J. Sahakian, Trevor W. Robbins, Lena Palaniyappan, Jie Zhang

**Author notes:** **Correspondence should be addressed to: Jie Zhang**, Institute of Science and Technology for Brain Inspired Intelligence, Fudan University, Shanghai, PR China, **Lena Palaniyappan**, Department of Psychiatry and Robarts Research Institute, University of Western Ontario, London, Ontario, Canada. These authors contributed equally to the work (Gechang Yu, Xinran Wu, Zhaowen Liu, Benjamin Becker, Kai Zhang).

## Abstract

Overlap of brain changes across mental disorders has reinforced transdiagnostic models. However, the developmental basis for this overlap is unclear as are neural differences among internalizing, externalizing and thought disorders. These issues are critical to inform the theoretical framework for hierarchical transdiagnostic psychiatric taxonomy. We examined cortical thickness (CT) difference between healthy controls (n=4041) and patients with externalizing (n=1182), internalizing (n=1959) and thought (n=347) disorders in preadolescents (9-10 years) from the Adolescent Brain and Cognitive Development Study using linear mixed models. Genome-wide association analysis and cell type specificity analysis were performed on regional CT across 4,716 unrelated European youth. We found that youth with externalizing or internalizing disorders, but not thought disorders, exhibited significantly thicker cortex than controls. Externalizing and internalizing disorders shared thicker CT in left pars opercularis and caudal middle frontal gyrus related to lower cognitive performance. Somatosensory and primary auditory cortex were uniquely affected in externalizing disorders; primary motor cortex and higher-order visual association areas were uniquely affected in internalizing disorders. Only group of externalizing disorders demonstrated significant CT increase than controls at 2-year follow-up and decelerated cortical thinning from 10 to 12 years old. At genetic level, genes associated with CT in common and disorders-specific regions were also implicated in related diagnostic families. Microglia were the cell-type associated with CT for both externalizing/internalizing while dopaminergic/glutamatergic/GABAergic cells related only to externalizing-specific regions. These results showed that distinct anatomical trajectories relevant to internalizing/externalizing phenotypes may result from unique genetic and cell-type changes, but these occur in the background of significantly shared morphological variance.

## Introduction

During the past two decades, conventional diagnostic categories of mental disorders have been increasingly challenged on the basis of unclear boundaries, overlapping symptoms [1, 2] as well as high comorbidity [3, 4]. Further, there is strong evidence of shared genetic risk [5-7] among various disorders, implying that the pathophysiology underlying these disorders may not be unique to each. While a number of shared features of neural dysfunction across mental disorders have been reported [8-10], any evidence for unique developmental pathways underlying these pathological endpoints has not been conclusively demonstrated.

Emerging transdiagnostic or dimensional frameworks, such as Research Domain Criteria (RDoC) [11] and The Hierarchical Taxonomy of Psychopathology (HiTOP) [12], have been proposed to transcend the limitations of conventional diagnostic categories of mental disorders. Based on accumulating evidence the HiTOP proposes that single mental disorders could be classified into three broad diagnostic families: externalizing disorders (inattention, aggressive and disruptive behavior), internalizing disorders (depression, anxiety and fear) and thought disorders (delusions, hallucinations and obsessions). Accordingly, a growing literature [13-16] has focused on mapping neural correlates for general psychopathology (‘p factor’), reflecting an overarching susceptibility to any mental disorder [17, 18], and several specific low-order broad diagnostic families (e.g., externalizing, internalizing and thought disorders). This hierarchical framework of mental disorders removes unclear boundaries between single mental disorders by grouping disorders with related symptoms into broad diagnostic families, which may further contribute to determining the underlying pathological dimensions on the neural, genetic and phenotype level [19, 20].

During childhood and adolescence, the brain undergoes major developmental changes in cortical morphology. One of the most fundamental neurodevelopmental changes involves cortical thickness (CT) which undergoes accelerated thinning during early adolescence compared with early childhood and adulthood [21]. This process is presumably driven by increased intracortical myelination [22] as well as synaptic pruning [23]. The disruption of CT has been linked to various psychopathology [24, 25] and impaired cognitive performance [26]. Most mental disorders originate in this developmental period [3, 27], highlighting the importance of examining CT as a marker of preadolescent mental health. Prior studies with limited samples recruited on the basis of traditional diagnostic categories have so far provided conflicting results with respect to the CT changes in this age group [24, 28].

In the current study, we combined case-control analyses with a hierarchical framework of mental disorders to examine CT alterations among three broad diagnostic families (externalizing, internalizing and thought disorders). We hypothesized that some of the topography of anatomical distribution of CT aberrations will be shared among the three diagnostic families, reflecting a shared mechanistic basis, while each diagnostic family will also be related to its own unique pattern of CT changes both at baseline and over time. To eliminate confounding effects introduced by high comorbidity among three diagnostic families [29] and facilitate the determination of disorders-specific alterations, our case-control analyses only used non-comorbid (i.e., “pure”) patients. Given that the neurodevelopmental patterns of CT during childhood and adolescence are genetically regulated [30, 31], we also undertook Genome-wide association study (GWAS) to locate the genetic variants associated with regional CT changes in all unrelated European youth. In the longitudinal analysis, we used 2-year follow-up data (n=6571) to examine whether regions with significant CT increase identified at baseline would persist at 2-year follow-up and whether three diagnostic families had distinct longitudinal changes in regional CT across a 2-year-age span, compared with healthy control (HC) group.

## Methods and materials

### Participants

Participants were preadolescents aged 9-10 years (n=11,878) recruited from 22 research sites across the USA from the Adolescent Brain Cognitive Development (ABCD) Study^®^ (Release3.0, November 2020). This longitudinal multisite population-representative cohort provides comprehensive clinical, behavioral, cognitive, and multimodal neuroimaging data from the baseline, 1-year and 2-year follow-up (12 years old) assessment. Inclusion/exclusion criteria for samples in all analyses were illustrated in Fig. S1-5.

### Measures

#### Definitions of diagnostic families

Single mental disorder diagnoses were determined using parent or guardian ratings in the computerized Kiddie Schedule for Affective Disorders and Schizophrenia (KSADS) based on DSM-5 criteria [32]. For the present analyses life-time (past or present) diagnoses of the 18 single mental disorders (Fig. 1A and Supplemental Table S1) used in our analyses were determined. Based on the definitions of broad diagnostic families in recent studies [13, 16, 29], three broad diagnostic families in our analyses (Fig. 1A) were determined by grouping 18 single mental disorders into externalizing disorders, internalizing disorders, thought disorders. As expected there was a high comorbidity among the three broad diagnostic families (Fig. 1B and Table 1). To eliminate confounding effects introduced by comorbidities, participants with comorbidities outside their primary diagnostic families were excluded from the following analyses. Healthy controls (Table 1) were those who did not meet any mental disorders diagnosis criteria of KSADS (including those disorders that are not included in three broad diagnostic families, such as Eating Disorders and Sleep Problems, where both unspecified or other specified disorders were covered). Demographics for patients of each single mental disorder were shown in Supplemental Table S1.

**Fig. 1.**
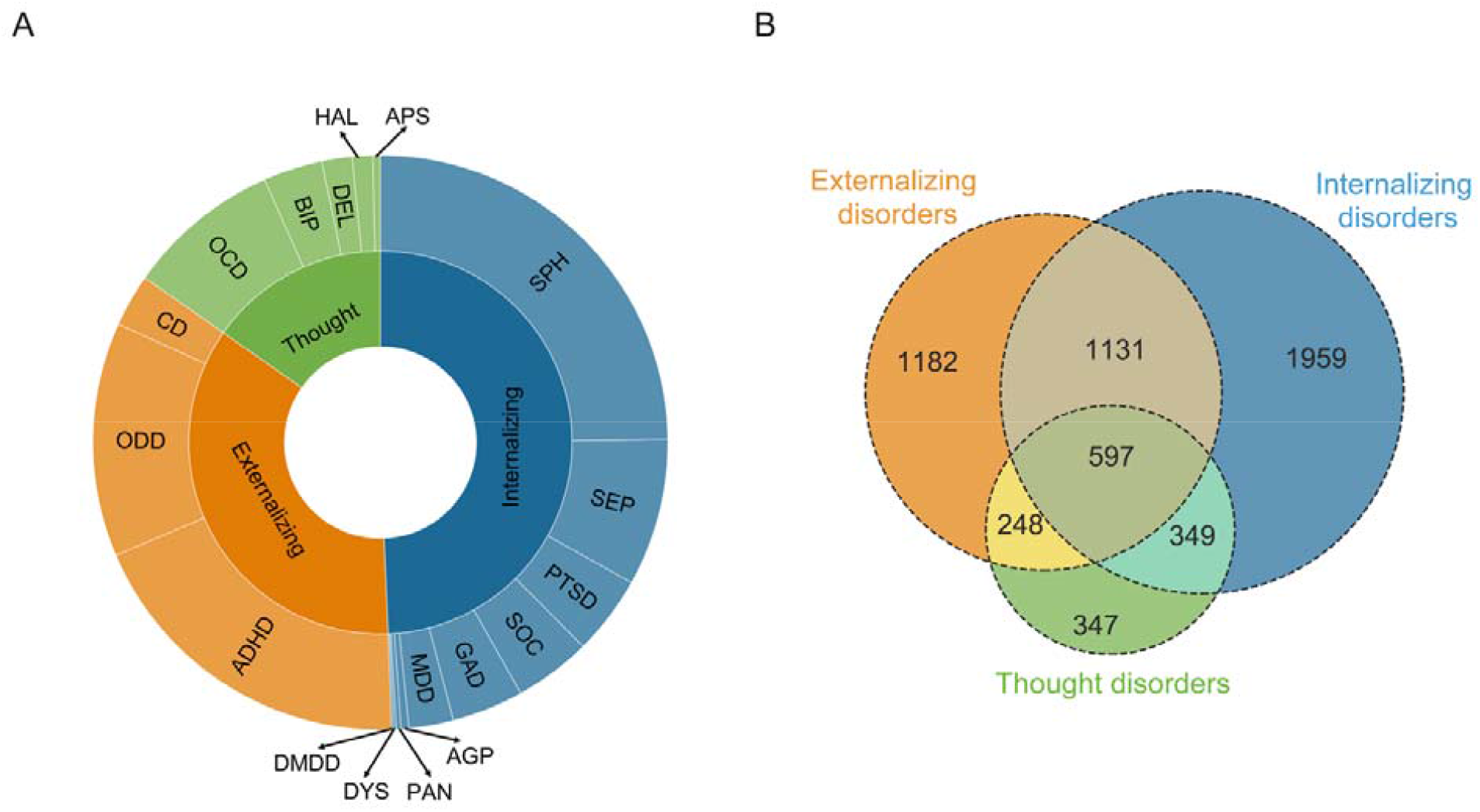
Components and comorbidity of externalizing, internalizing and thought disorders. **A** 18 mental disorders (outer circle) were classified into three broad diagnostic families (inner circle), i.e., externalizing, internalizing and thought disorders. **B** Venn diagram depicts the large overlap among three diagnostic families. Pure subsets of three diagnostic families: externalizing disorders, orange; internalizing disorders, blue; thought disorders, green. Abbreviations: ADHD= Attention Deficit/Hyperactivity disorder, CD=Conduct Disorder, ODD=Oppositional Defiant Disorder, MDD=Major Depressive Disorder, GAD=Generalized Anxiety disorder, SOC=Social Anxiety Disorder, SEP=Separation Anxiety Disorder, PTSD=Post-Traumatic Stress Disorder, AGP=Agoraphobia, SPH=Specific Phobia, PAN=Panic Disorder, DYS=Dysthymia, DMDD=Disruptive Mood Dysregulation Disorder, BIP= Bipolar Disorder, OCD=Obsessive-Compulsive Disorder, DEL= Delusions, HAL=Hallucinations, APS= Associated Psychotic Symptoms.

**Table 1.** Demographics of three subsets of broad diagnostic families without comorbidity.

|  | Externalizing disorders without comorbidity | Internalizing disorders without comorbidity | Thought disorders without comorbidity | Healthy controls |
| --- | --- | --- | --- | --- |
| N | 1182 | 1959 | 347 | 4041 |
| Age (months) | 119.18(7.39) | 119.24(7.54) | 118.79(7.42) | 119(7.48) |
| Sex (male) | 804(68.02%) | 896(45.74%) | 182(52.45%) | 1885(46.65%) |
| Race |  |  |  |  |
| White | 661(55.92%) | 1069(54.57% ) | 150(43.23%) | 2029(50.21%) |
| Black | 179(15.14%) | 258(13.17%) | 79(22.77%) | 604(14.95%) |
| Hispanic | 204(17.26%) | 400(20.42%) | 72(20.75%) | 890(22.02%) |
| Asian | 12(1.02%) | 36(1.84%) | 6(1.73%) | 112(2.77%) |
| Others/Mixed | 126(10.66%) | 196(10.01%) | 40(11.53%) | 406(10.05%) |

#### Child Behavior Checklist (CBCL)

The Child Behavior Checklist (CBCL), completed by the child’s parent or caregiver, is widely used to assess emotional and behavioral problems in the children. In the current analyses, we used raw scores of 20 CBCL scales (details were described in Supplemental materials) from the baseline (n=11,870) and from the 2-year follow-up (n=6,571).

#### NIH Toolbox^®^ cognition measures

NIH Toolbox^®^ consists of seven tasks, measuring executive function, episodic memory, working memory, information processing and language abilities and three summary scores including crystal intelligence, fluid intelligence and total intelligence. Baseline data included 11,878 individuals while 2-year follow-up data included 6,571 individuals with only 6 cognition scores. Details of cognitive measures were listed in Supplemental Table S3.

#### Structural image acquisition and quality control

Participants completed a high-resolution T1-weighted structural MRI scan (1-mm isotropic voxels) on 3T scanners (Siemens Prisma, General Electric MR 750, Philips). Structural MRI data processing were completed using FreeSurfer version 5.3.0 according to standardized processing pipelines [33]. All scans underwent radiological review to identify incidental findings. Participants who did not pass visual inspection of T1 images and FreeSurfer quality control [34] (imgincl_t1w_include==1) were excluded from the neuroimaging analyses. The current study used post-processed cortical thickness data mapped to 34 cortical parcellations per hemisphere (68 total regions of interest) based on the Desikan-Killiany Atlas [35].

#### Genetic data

Using the IMPUTE2 software and the imputation reference set obtained from the Phase 3 of 1000 Genomes Project, we implemented genotype imputation on high-quality genotyped data consisting of 11,099 individuals and 516,598 SNPs (single nucleotide polymorphism). Post-imputation quality control excluded individuals with >10% missing rate and SNPs with imputation info score<0.7, >10% missing rate, MAF (Minor Allele Frequency) <0.5%, or out of Hardy-Weinberg equilibrium violation (p>10^−6^). We then derived genetic relatedness (kinship coefficient) using plink v2.0. GWAS and its subsequent analysis only included European ancestry (genetic ancestry factor of European>0.9) and genetically unrelated (kinship coefficient<0.125) participants, containing 4,933 individuals and 8,498,283 SNPs. In order to correct for population stratification, genetic Principal Component Analysis (PCA) were performed on these unrelated European individuals to derive first 10 genetic principal components (PCs).

### Statistical Analyses

#### Case-control analysis and ANOVA

We implemented linear mixed models (LMM) using the R *lme4* [36] package to examine difference in CT among the 3 “pure” diagnostic families in contrast to HC. In LMM, CT was dependent variable and group (HC defined as 0 and patients defined as 1) was independent variable. All LMM also included random effects for family nested within acquisition site and fixed-effects covariates for age, sex, race (White, Black, Hispanic, Asian, Others/Mixed), parental marital status, pubertal level, parental education, body max index (BMI) and total intracranial volume. False Discovery Rate (FDR) were used in all analyses for multiple comparisons. We also examined the CT alterations in three diagnostic families encompassing comorbid cases (See Supplemental materials, Fig. S6A-C). Then we calculated the Pearson correlation among the whole brain T-maps of three diagnostic families to examine the similarity of CT alterations between diagnostic families. To examine the difference in CT among four groups (externalizing, internalizing and thought disorders and healthy preadolescents), we also undertook an Analysis of Variance (ANOVA) after regressing out the same covariates using LMM. Post hoc analysis using Tukey HSD Test was also performed to compare the four groups.

A common alteration in three diagnostic families was defined as a significant difference (P_FDR_<0.05) in CT between any patient group and healthy control (HC) group and shared by at least 2 patient groups. A disorders-specific alteration (e.g., externalizing-specific) is defined by 1) a significant difference between one patient group and HC group, and 2) a nonsignificant difference (P_FDR_>0.05) between each of the other two patient groups and HC group. This definition was in line with our prior work [37]. Notably, this definition of disorder specificity does not indicate exclusivity (i.e., absence of a corresponding change in another patient group), but only indicates that a change with an effect size sufficient for detection at group level is specific to a group.

#### Correlation and longitudinal analysis

We performed correlation analyses between CT in common and disorders-specific regions and total CBCL scores as well as cognition scores at baseline in the whole sample using the same LMM. We also restricted the above analyses to externalizing or internalizing disorders groups to examine whether patient groups exhibited distinct associations (Supplemental materials). We further performed correlation analyses between baseline CT and the CBCL and cognitive scores at 2-year follow-up to examine if baseline cortical thickness could predict the development of behavioral problems and cognition.

We used 2-year follow-up CT data (575 externalizing disorders, 913 internalizing disorders, 169 thought disorders and 1881 HCs) to evaluate the longitudinal changes in the disorders groups: 1. we performed case-control study for all 3 diagnostic families to see if baseline CT alterations would persist after 2 years. 2. using LMM, we compared longitudinal CT changes from 10 to 12 years old between three diagnostic families and HC group to identify abnormal cortical thinning. The LMM additionally included random intercepts defining subject as random factor, time variable (baseline defined as 0) and group-by-time interactions, which measured the case-control differences in cortical thinning from 10 to 12 years old. All reported comparisons were FDR corrected for the number of regions of interest (ROIs).

### Genome-wide association study (GWAS)

We performed GWAS to examine the genetic variants underlying regional CT using plink v2.0. On the premise of additive genetic effects, general linear regression models were fitted to determine the association between common and disorders-specific CT alterations and allele dosages of SNPs in genetically unrelated European-ancestry preadolescents who passed structural image quality control (n=4,716). Sex, age, mean cortical thickness, 10 PCs and study sites were included as covariates. Then SNPs were annotated and mapped to genes (See supplemental materials) by FUMA [38] online platform (version 1.3.6a), which is an integrative tool for functional mapping and annotation of genetic associations.

### Gene set enrichment analysis (GSEA)

Genes identified by FUMA were merged separately across externalizing-specific, internalizing-specific and common regions. To gain insight into biological functions and pathways of externalizing-specific, internalizing-specific and common genes, GSEA was performed to test if these genes are overrepresented in pre-defined gene sets obtained from the Molecular Signatures Database [39] and GWAS catalog [40]. All genes were set as background genes. FDR correction was performed per gene set by FUMA. Other parameters in these analyses were set as default.

### Cell type specificity analysis (CTSA)

In order to examine the associations between regional CT and specific cell types through gene expression, we performed CTSA [41] implemented in FUMA, using 7 single-cell RNA sequencing datasets from human brain tissue (Supplemental Table S4) and associations between genes and regional CT. We used FDR correction for multiple testing in per dataset to identify significantly associated cell types.

## Results

### Common and unique cortical thickness alterations

Of a total of 68 regions, CT was significantly higher in internalizing (13 regions) and externalizing (9 regions) disorders compared with healthy youth. No regions were significantly altered in either direction in thought disorders (after FDR correction). CT increases converged across externalizing and internalizing disorders in 2 left lateral frontal regions (Fig. 2A-B): left pars opercularis and left caudal middle frontal gyrus (MFG). Externalizing-specific changes were also left-lateralized, being localized to the left transverse temporal gyrus, left superior temporal gyrus (STG), left postcentral gyrus, left superior frontal gyrus (SFG) and left isthmus of the cingulate cortex. Internalizing-specific changes were localized to bilateral precentral gyrus, right inferior temporal gyrus (ITG) and left fusiform gyrus.

**Fig. 2.**
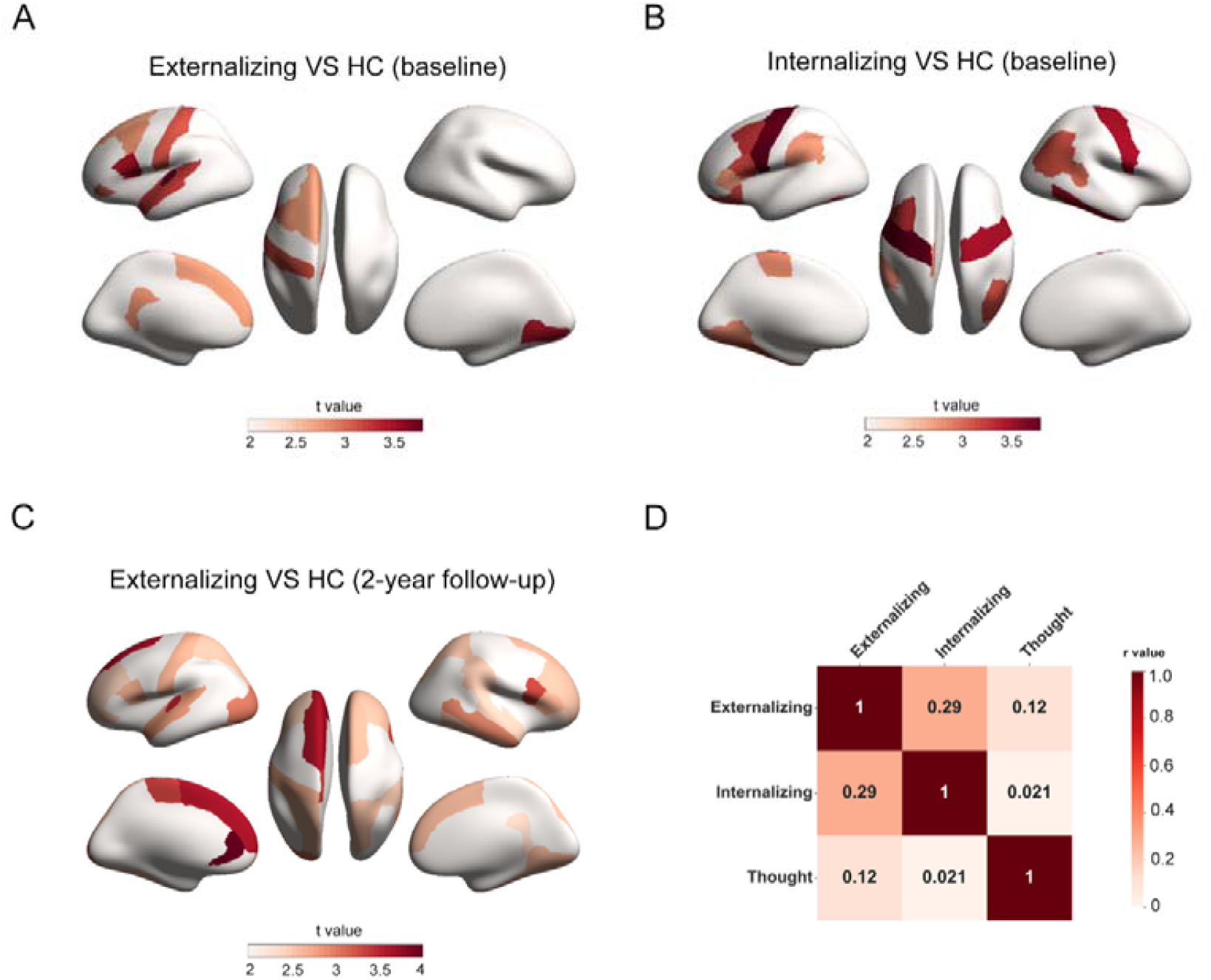
Regions with significant (P_FDR_<0.05) thickness alterations at baseline and 2-year follow-up. Regions with significant (P_FDR_<0.05) thickness alterations at baseline in **A** externalizing disorders and **B** internalizing disorders and **C** at 2-year follow-up in externalizing disorders. **D** correlations among baseline T-maps of three diagnostic families. The colorbars in **A, B** and **C** represent the t value of the regression coefficient of group variable from LMM. The colorbar in **D** represents Pearson correlation coefficient between baseline T-maps of three diagnostic families. Abbreviations: Externalizing=externalizing disorders, Internalizing=internalizing disorders, Thought=thought disorders, HC=healthy control.

The whole brain T-map (Fig. 2D) of externalizing disorders vs. HC contrast had a moderate level of spatial correlation with that of internalizing disorders vs. HC (r=0.294, p=0.015). Interestingly, the T map of thought disorders contrast did not correlate with the other two disorders.

ANOVA contrasting the four groups (externalizing, internalizing and thought disorders and HC) identified five significant (P_FDR_<0.05) regions, including left pars opercularis, bilateral precentral gyrus, left caudal MFG and left transverse temporal gyrus (Supplemental Table S5). Post hoc analyses of a direct contrast revealed that CT of the left pars opercularis and caudal MFG (common regions) was thicker in both externalizing and internalizing disorders compared with HC, which was consistent with independent contrasts results. CT in left transverse temporal gyrus (externalizing-specific region) was also thicker in externalizing disorders than in HC. Notably, CT of bilateral precentral gyrus (internalizing-specific regions) was thicker in internalizing disorders than in both healthy youth and externalizing disorders.

### Association between altered cortical thickness and behavioral symptoms and cognition

For associations between CBCL scores and CT in regions with a significant diagnostic effect (Fig. 3), only CT in the left isthmus of cingulate cortex positively correlated with CBCL Externalizing Problems (p= 0.002, t=3.032). No associations with internalizing-specific or common regions survived FDR correction. Regarding cognitive performance (Fig. 3), List Sorting Working Memory score had notable associations with transdiagnostically affected, externalizing-specific and internalizing-specific regions. For regions affected transdiagnostically, CT in the left pars opercularis was higher in subjects with lower List Sorting Working Memory score (p=4.6×10^−5^, t=-4.0775), Oral Reading Recognition score (p=2.7×10^−4^, t=-3.649), crystal intelligence (p=0.002, t=-3.059) and total intelligence (p=0.001, t=-3.269). CT in the left caudal middle frontal gyrus was also higher in subjects with lower List Sorting Working Memory score (p=8.6×10^−4^, t=-3.333).

**Fig. 3.**
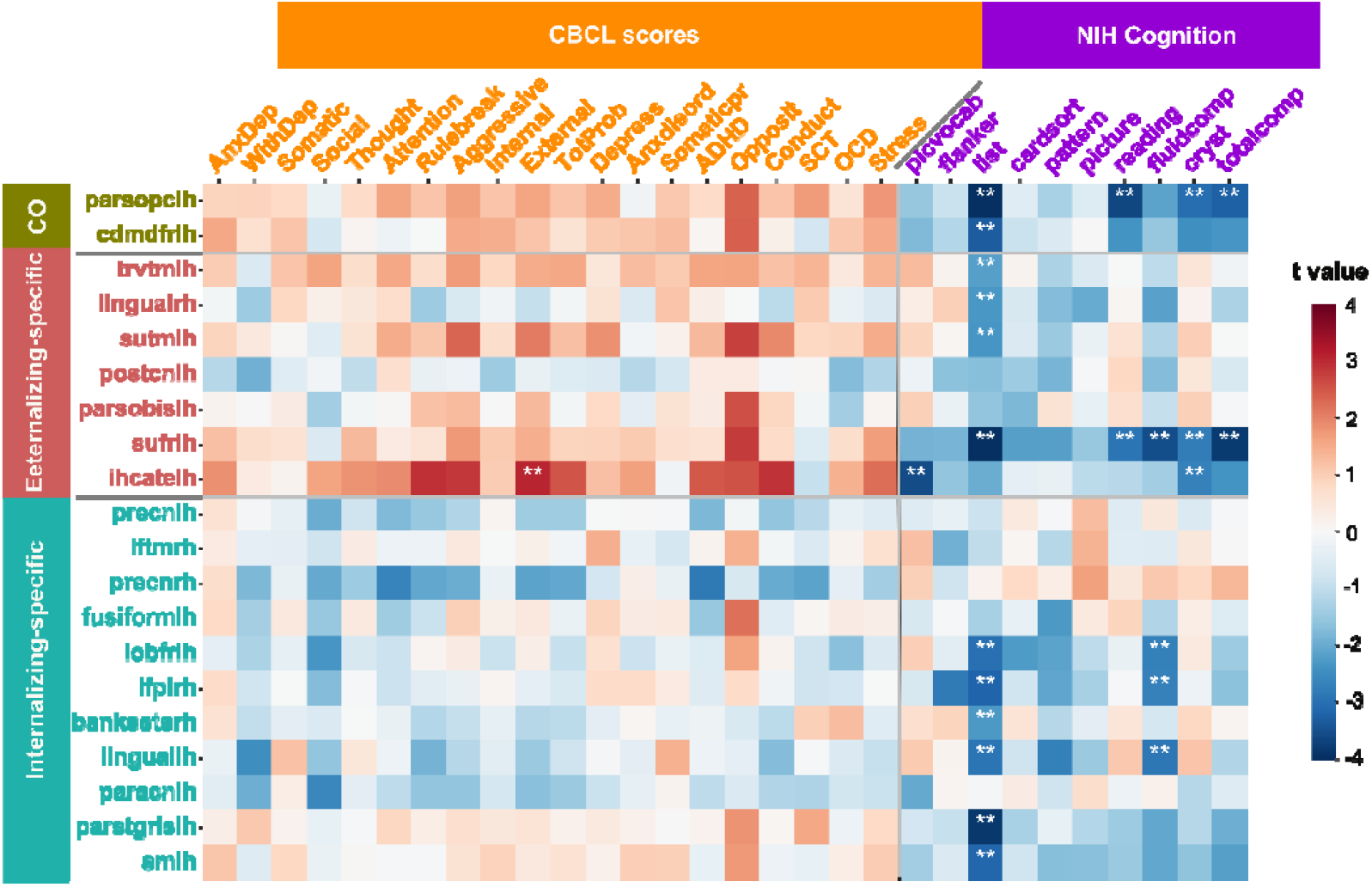
Associations between baseline CT in the Common (CO), Externalizing-specific and Internalizing-specific regions, baseline CBCL subscales, and NIH cognition scores. The colorbar represents the t value of the regression coefficient from LMM. Two asterisks (**) indicate P_FDR_<0.05. Abbreviations: parsopclh=left pars opercularis, cdmdfrlh=left caudal middle frontal, trvtmlh=left transverse temporal, lingualrh=right lingual, sutmlh=left superior temporal, postcnlh=left postcentral, parsobislh=left pars orbitalis, sufrlh=left superior frontal, ihcatelh=left isthmus of cingulate cortex, precnlh=left precentral, iftmrh=right inferior temporal, precnrh=right prencentral, fusiformlh=left fusiform, lobfrlh=left lateral orbitofrontal, ifplrh=right inferior parietal lobule, banksstsrh=right banks of superior temporal sulcus, linguallh=left lingual, paracnlh=left paracentral, parstgrislh=left pars triangularis, smlh=left supramarginal, AnxDep=Anxious/Depressed, WithDep=Withdrawn/Depressed, Somatic=Somatic Complaints, Social=Social Problems, Thought=Thought Problems, Attention=Attention Problems, Rulebreak= Rule-Breaking Behavior, Aggressive=Aggressive Behavior, Internal=Internalizing Problems, External=Externalizing Problems, TotProb=Total Problems, Anxdisord=Anxiety disorders, Somaticpr=Somatic Problems, SCT= Sluggish Cognitive Tempo, Opposite= Oppositional Defiant Problems, Conduct=Conduct Problems, OCD=Obsessive-Compulsive Problems, Stress=Stress Problems, ADHD=Attention Deficit/Hyperactivity Problems, picvocab=Picture Vocabulary, flanker=Flanker Inhibitory Control and Attention, list= List Sorting Working Memory, cardsort= Dimensional Change Card Sort, pattern=Pattern Comparison Processing Speed, Picture=Picture Sequence Memory, reading=Oral Reading Recognition, fluidcomp=fluid composite, cryst= crystallized composite, totalcomp=total composite.

Other cognitive scores also related to CT in regions specifically affected in externalizing and internalizing disorders. In general all relationships were negative correlations, suggesting higher CT in these regions related to poor cognitive performance in the affected domains. With respect to the 2-year follow-up CBCL scores (Fig. S7), only externalizing-specific regions showed significant associations. Thicker CT of the left isthmus of the cingulate cortex predicted higher CBCL scores related to externalizing problems. CT of this region correlated positively with CBCL scales of externalizing disorders at baseline and after two years, suggesting it is a stable predictor of externalizing behaviors or symptoms. Further, these associations cut across diagnostic families and extend to heathy controls, indicating a generalized and continuous relationship between CT and cognitive measures, akin to the p factor.

### Longitudinal analyses

For CT alterations at 12 years old (2-year follow-up), externalizing disorders showed significantly thicker CT in 26 regions than controls (Fig. 2C), 7 of which (left transverse temporal gyrus, left STG, left postcentral gyrus, left SFG, right lingual gyrus, left pars orbitalis and left pars opercularis) were overlapped with baseline results. No regions survived FDR correction in internalizing and thought disorders. As to longitudinal changes in regional CT (i.e., within subject changes from 10 to 12 years old), externalizing, internalizing and thought disorders and HC all showed pervasive cortical thinning from 10 to 12 years old (Fig. S8A-D). However, only externalizing disorders showed decelerated cortical thinning compared with HC group (Fig. 4A-B, including right pars opercularis, right parahippocampal gyrus, bilateral insula, left fusiform gyrus and right transverse temporal gyrus).

**Fig. 4.**
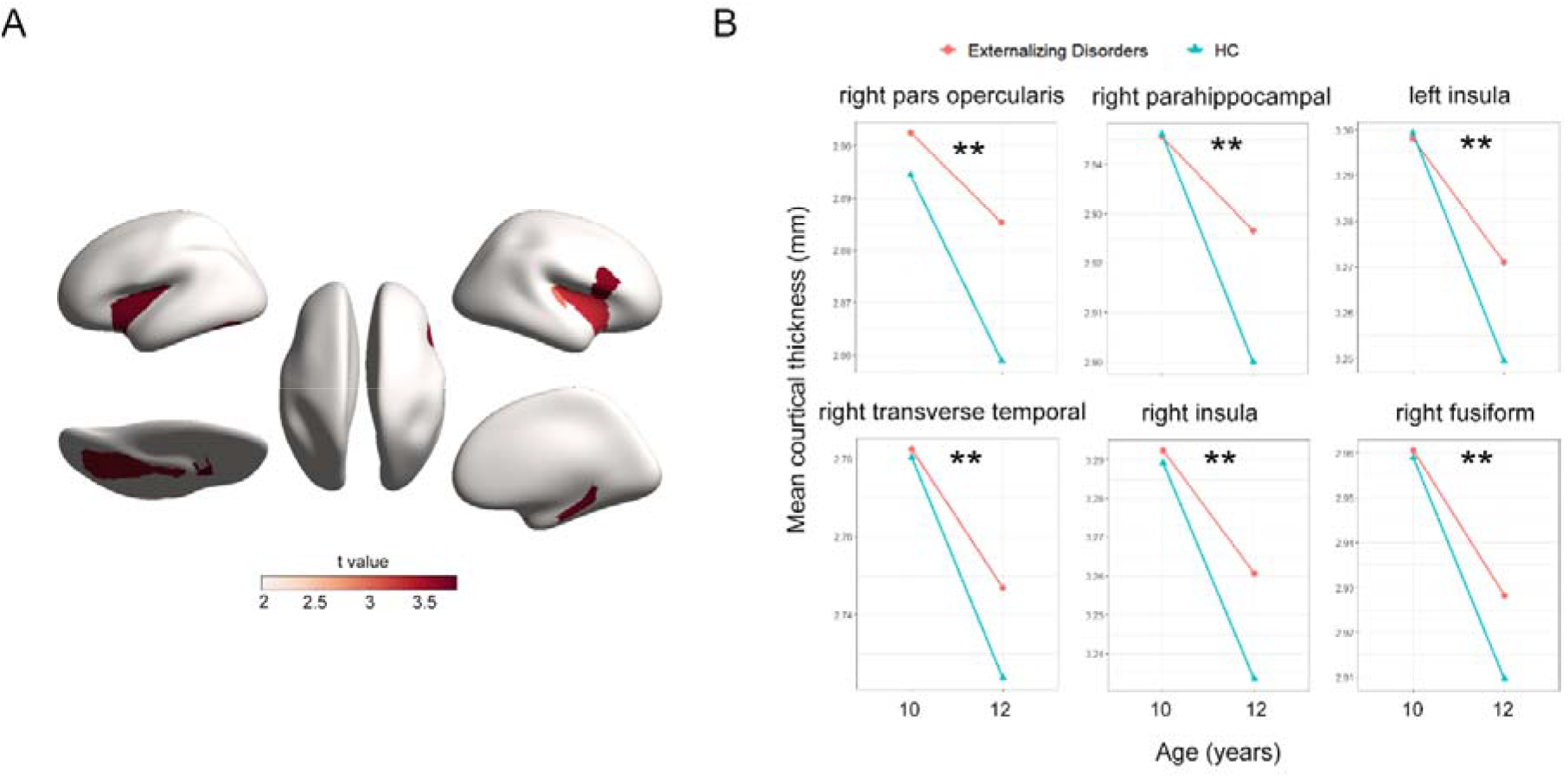
Regions with significantly decelerated cortical thinning from 10 to 12 years old in externalizing disorders compared with HC group. **A** Regions with significantly (P_FDR_<0.05) decelerated cortical thinning from 10 to 12 years old in externalizing disorders (compared with HC). The colorbar in **A** represents the t value of the regression coefficient of group-by-time interaction from LMM. **B** Mean cortical thickness of above regions at 10 and 12 years old in externalizing disorders and HC. Two asterisks (**) indicate P_FDR_<0.05.

### GWAS, GSEA and CTSA

We performed GWAS of CT of common and disorders-specific regions (all together 20 significant ROIs) using 4,716 European-ancestry unrelated individuals whose structural image passed quality control. Under the classic genome-wide threshold of P<□5□×□10^−8^, there were 8 regions with significant associations (Supplemental Table S6-13), including one common region (left pars opercularis), two externalizing-specific regions (left postcentral gyrus and right lingual gyrus) and five internalizing-specific regions (right banks of superior temporal sulcus, right inferior parietal lobule, left paracentral gyrus and bilateral precentral gyrus). The strongest association with regional CT were observed for rs76289836 at 11q13.4 in the right banks of superior temporal sulcus (STS, p=6.4×□10^−20^). Although this locus has not been previously reported, its nearest gene IGF2 (insulin growth factor 2), has been linked to anxiety [42] and PTSD [43]. rs2033939 at 15q14, which was associated with CT in postcentral gyrus (p=2.7×□10^−15^), has also been associated with CT in postcentral gyrus in three previous GWAS [44-46]. Another variant related to CT in postcentral gyrus, rs1080066 at 15q14 (p=5.6×□10^−15^), has been highlighted in two recent GWAS [46, 47], where it was associated with cortical area (CA) in precentral gyrus.

FUMA identified 112, 131 and 1172 genes associated with CT of common, externalizing-specific and internalizing-specific regions (Supplemental Table S14-16), respectively. We found that genes associated with CT of common, externalizing-specific and internalizing-specific regions have also been linked to the pathology of the related diagnostic families. For example, for common regions like left pars opercularis, the identified genes were implicated in either externalizing or internalizing disorders: ADGRL3 (formerly LPHN3, latrophilin subfamily of G-Protein-Coupled Receptor), near rs58251292 (p= 2.61×10^−9^), is strongly linked with ADHD [48, 49] in a large body of literature while Slit3, including rs142753275 (p= 2.75×10^−9^), has been associated with MDD [50]. GSEA also showed that genes related to CT of left pars opercularis were significantly associated with internalizing symptoms like “facial emotion recognition of sad faces” (p=5.28×10^−22^), “response to cognitive-behavioral therapy in anxiety and MDD” (p=1.01×10^−8^), and externalizing symptoms like “impulsivity” (p=2.54×10^−4^), see Supplemental Table S17-19.

Moreover, in externalizing-specific regions like left postcentral, rs551123675 and rs529206008, which were in LD with rs114949000 (p=7.85×10^−9^), were identified, located in VDR (Vitamin D Receptor) that have been reported to be related to ADHD [51]. In internalizing-specific regions like the right banks of STS, rs531970391, which is in LD with rs9996981 (p=9.94×10^−20^), located in FGF2 (fibroblast growth factor-2), has been implicated in anxiety and depression [52, 53]. Other genes linked to CT in this region like HTR1B and GRM8, have also been implicated in internalizing disorders [54, 55]. GSEA showed that genes associated with internalizing-specific regions were involved in neurogenesis, neuron death and differentiation.

CTSA identified shared cell types across externalizing and internalizing disorders and also disorders-specific cell types (Supplemental Table S20). Three internalizing-specific regions (right precentral, left pars triangularis and right ITG) and one externalizing-specific region (left postcentral) showed significant associations with microglia. CT in left postcentral was also associated with Ex6a (excitatory neurons), Glut3 and Glut4 (glutamatergic neurons). For other externalizing-specific regions, genetic variants for CT in the left SFG aggregated on exPFC1, exPFC2 (excitatory glutamatergic neurons from the prefrontal cortex), exCA1 and exCA3 (excitatory pyramidal neurons in the hippocampal Cornu Ammonis) while those for CT in the left isthmus of cingulate cortex aggregated on DA1 (dopaminergic neurons) and NbGaba (GABAergic neuroblasts). For common regions, CT in left pars opercularis is related to Nprog (neuronal progenitors) and ProgM (medial progenitors).

## Discussion

This cross-sectional and longitudinal study identified regions of increased CT shared by externalizing and internalizing disorders and regions unique to each of them in a large preadolescent sample consisting of 11,878 subjects. Previous findings from smaller samples, limited by comorbidities among single mental disorders and broad diagnostic families, have been inconsistent to date. Some [13, 24] have found reduced CT in internalizing/externalizing disorders or associated with internalizing/externalizing symptoms while others had opposite findings [24, 28, 56]. Our current results demonstrated increased CT characterizes in preadolescents with externalizing disorders as well as internalizing disorders. Our study provides an interesting insight when seen in conjunction with the adult sample (median age 45) of *Romer et al* [13] who reported notable cortical thinning in relation to the general psychopathology (‘p’ factor) as well as highly overlapping patterns across the three diagnostic families examined here. CT is one of the structural indices that is highly sensitive to age-related changes [57]. The developmental trajectory of CT in relation to ‘p’ factor is likely to be one of a leftward developmental shift, characterized by increased thickness due to deficient age-appropriate reduction in preadolescence, but accelerated thinning in adulthood due to excessive age-related loss [58, 59]. Alternatively, different mechanisms may be at play at different age groups; with lack of intracortical myelination [22] contributing to p factor in preadolescence; with dendritic spine reduction/synaptic pruning or increased myelination [58, 60] contributing to dysregulations in adulthood. It is also possible that changes during later adulthood are largely compensatory in nature, secondary to the early developmental deficits, though a convincing test of this structural compensation hypothesis is still lacking [61]. We noted no significant difference between thought disorders and HC, likely due to the smaller sample size, or larger shared variance between thought disorders and the HC, compared to the other 2 diagnostic families.

CT aberrations in the pars opercularis (BA44) of the left IFG and left caudal MFG were common to both externalizing and internalizing disorders. The pars opercularis, often seen as the site of Broca’s area, is not only involved in inhibitory control [62], but also plays a critical role in empathy [63], impairments of which are relate to externalizing disorders like ADHD [64]. Pars opercularis also located in the ventrolateral prefrontal cortex (VLPFC) that is involved in emotional regulation through direct projections to the ventral medial prefrontal cortex-amygdala pathway [65]. Thicker pars opercularis may result in impairment in emotional regulation, which is related with internalizing disorders like trait anxiety [66]. The caudal MFG corresponds to the dorsolateral prefrontal cortex (DLPFC), which plays an important role in a range of executive functions such as cognitive flexibility [67] and working memory [68]. Abnormalities of DLPFC may undermine the top-down cognitive control, which are implicated in both externalizing [69, 70] and internalizing disorders [71].

Externalizing and internalizing disorders also exhibited distinct patterns of CT increase. Interestingly, externalizing-specific regions included left postcentral gyrus responsible for somatosensory processing, while internalizing-specific regions included bilateral precentral gyrus which is primary motor cortex. ANOVA results also showed that CT in bilateral precentral gyrus was thicker in internalizing disorders than in externalizing disorders. Abnormalities of postcentral gyrus have been linked to externalizing disorders or symptoms [25, 72], which are likely to result in dysfunctions of somatosensory. Previous literatures have associated defects of somatosensory regions with ADHD [73]. Internalizing symptoms like anxiety could affect perceptual-motor performance [74]. Children with depression or anxiety disorders often exhibit poor motor skills and performance [75]. What’s more, psychomotor retardation is one of core symptoms of major depressive disorder (MDD). Therefore, alterations of cortical morphology in the precentral gyrus found in internalizing disorders like anxiety disorders [24] and MDD [76], may lead to impaired motor abilities and psychomotor retardation. Furthermore, a burgeoning body of literature [77-79] has focused on the role of somatosensory-motor network in psychopathology.

For visual and auditory cortices, we also found distinct alterations in externalizing and internalizing disorders. Externalizing patients demonstrated alterations more in primary auditory areas such as the STG and Heschl’s gyrus, while internalizing patients demonstrated alterations more in higher order visual association areas like the fusiform gyrus and ITG, which is involved in the higher order visual processing. Hypersensitivity or hyposensitivity to auditory stimulus are also common symptoms in ADHD. Functional alterations of STG and Heschl’s gyrus have been linked with ADHD [80] and conduct disorder [81]. The fusiform gyrus and ITG plays an important role in facial recognition and perception [82], abnormalities of which have been associated with internalizing disorders like depression [83]. Adolescents with recurrent depression also showed decreased surface area of the left fusiform gyrus [83]. ITG is also a key part of the ventral visual pathway [84], linked with internalizing disorders [85, 86]. Finally, associations of the T map (Fig. 2C) among three diagnostic families showed that the alteration patterns in externalizing and internalizing disorders were only slightly correlated, indicating different neural mechanism underlying externalizing and internalizing diagnostic family.

For 2-year follow-up (12 years old), only children with externalizing disorders displayed significantly thicker CT compared to controls, which is more widespread than baseline (with 7 regions overlapped with baseline results, including prefrontal regions like pars opercularis, pars orbitalis and SFG), validating the regions identified at baseline. Furthermore, among all 3 diagnostic families, only externalizing disorder group showed decelerated cortical thinning from 10 to 12 years old compared to controls. This is consistent with previous literatures showing reduced cortical thinning in externalizing disorders or symptoms in adolescence [25, 87, 88]. These results suggested that pathology related to CT alteration in externalizing disorders are relatively stable during early adolescence.

GWAS, GSEA, and CTSA identified common and unique genetic factors (genetic variants, genes, biological functions and cell types) underlying externalizing and internalizing disorders, providing genetic support for common and disorder-specific CT changes. Through GWAS, we detected the genes associated with CT in common (ADGRL3 and Slit3), externalizing-specific (VDR) and internalizing-specific (IGF2, FGF2, HTR1B and GRM8) regions, which are also implicated in the related diagnostic families. Genes associated with CT of one common region (left pars opercularis) were found to be enriched in both externalizing symptoms (“impulsivity”) and internalizing symptoms (“facial emotion recognition of sad faces” and “response to cognitive-behavioral therapy in anxiety and MDD”), further highlighting CT of left pars opercularis as a common biomarker of externalizing and internalizing disorders.

Moreover, CTSA identified both shared (microglia) and unique (glutamatergic neurons, dopaminergic neurons and GABAergic neuroblasts) cell types for different diagnostic families. Microglia were associated with CT in both externalizing-specific and internalizing-specific regions, suggesting that microglia may be a promising target for treatment of mental disorders. Microglia are the major immune cells of brain and also plays a role in synaptic plasticity, neurogenesis and memory [89, 90]. Previous studies have suggested that microglial dysfunction is associated with both externalizing disorders like ADHD [91] and internalizing disorders like depression [92, 93]. We found glutamatergic neurons, dopaminergic neurons and GABAergic neuroblasts were exclusively related with CT in externalizing-specific regions, and changes in glutamate/glutamine, dopamine neurotransmission pathway and GABA have been associated with ADHD [94-96]. In summary, our genetic results revealed that there are both common and unique genetic factors underlying externalizing and internalizing disorders just as there are common and unique neural factors.

The current study has several strengths. Firstly, ABCD is a multisite large-scale population-representative adolescent cohort with comprehensive psychopathology assessments. Diagnoses of over twenty adolescent disorders allowed us to explore neural mechanisms underlying broad diagnostic families. Secondly, our work is the first to combine a case-control study with HiTOP in a preadolescents cohort, thus also helping to more clearly demarcate boundaries between mental disorders. Furthermore, we only used non-comorbid patients, thus eliminating the interference from comorbidity among diagnostic families. Thirdly, longitudinal analysis further validated the significantly altered regions found at baseline and examined the abnormal longitudinal changes of CT in three diagnostic families. Finally, genetic data were also integrated into our study to further delineate the genetic factors influencing CT alterations. To our knowledge, it is the largest GWAS of preadolescent brain imaging phenotypes so far.

We also need to consider some limitations. No significant CT alterations between thought disorders and HC may be due to its small sample size (n=347). The follow-up data is only half of that at baseline, and results from longitudinal analyses also need to be further validated. Finally, the sample size in our GWAS analysis is smaller than those in traditional GWAS analysis, our genetic results therefore should be taken with caution and need further validations in larger samples.

In conclusion, the current study identified common and unique regional CT alterations in externalizing and internalizing disorders at baseline. Most thicker regions in externalizing disorders still persisted at 2-year follow-up, and regional decelerated cortical thinning from 10 to 12 years old was also found for externalizing disorders, suggesting CT alterations were more stable in externalizing disorders. We also performed GWAS, GSEA and CTSA and identified shared and unique genetic factors underlying externalizing and internalizing disorders. Microglia were the cell-type associated with CT for both externalizing and internalizing disorders while dopaminergic/glutamatergic/GABAergic cells related only to externalizing-specific regions. More importantly, our findings underscore the importance of searching for specificity of neural and genetic mechanisms underlying broad diagnostic families of mental disorders.

## Data Availability

GWAS summary statistics of regional CT of 20 ROIs could be downloaded on (https://drive.google.com/drive/folders/1a64gvTM5AQAYLdUw7GQxT-GQN1msXw3H?usp=sharing).

## Data Availability

GWAS summary statistics of regional CT of 20 ROIs could be downloaded on https://drive.google.com/drive/folders/1a64gvTM5AQAYLdUw7GQxT-GQN1msXw 3H?usp=sharing.

## Acknowledgments

JZ was supported by Shanghai Municipal Science and Technology Major Project (No.2018SHZDZX01) and ZJLab and NSFC 61973086. JF was supported by the 111 Project (No. B18015), the key project of Shanghai Science and Technology (No. 16JC1420402), National Key R&D Program of China (No. 2018YFC1312900), National Natural Science Foundation of China (NSFC 91630314). LP acknowledges salary support from the Tanna Schulich Chair of Neuroscience and Mental Health.

