## Supplemental materials for "Common and Disorders-Specific Cortical Thickness Alterations in Internalizing, Externalizing and Thought Disorders over a 2-year Period in the Preadolescents of the ABCD Study"

CBCL scores

The composite CBCL scores used in our study included eight syndrome scale scores (Anxious/Depressed, Withdrawn/Depressed, Somatic Complaints, Social Problems, Thought Problems, Thought Problems, Rule-Breaking Behavior, Aggressive Behavior), three summary scores (Internalizing Problems, Externalizing Problems and Total Problems), six DSM-oriented scale scores (Depressive Problems, Anxiety Problems, Somatic Problems, Attention Deficit/Hyperactivity Problems, Oppositional Defiant Problems and Conduct Problems) and three 2007 Scale Scores (Sluggish Cognitive Tempo, Obsessive-Compulsive Problems and Stress Problems).

SNP annotation and mapping

Genomic risk loci were defined using the FUMA [1] online platform (version 1.3.6a). Independent significant SNPs (IndSigSNPs) were defined as variants with a P-value< 5×10^-8^ and independent of other significant SNPs at r^2^<0.6. Lead SNPs were also identified as those independent from each other (r^2^<0.1). LD (Linkage Disequilibrium) blocks for IndSigSNPs were then constructed by tagging all SNPs with MAF (Minimum Allele Frequency)≥0.0005 and in LD (r^2^≥0.6) with at least one of the IndSigSNPs. The reference panel population was European of the 1000 Genomes Project Phase 3.

To further link these associated SNPs to genes, three strategies implemented by FUMA were employed: positional mapping, eQTL (expression quantitative trait loci) mapping and 3D Chromatin Interaction mapping. In addition, to combine cumulative effects of SNPs assigned to a gene, gene-based association analysis was performed using MAGMA [2] implemented in FUMA. SNPs were mapped to protein-coding genes if they are located within the genes. The gene-based P-value for each gene was calculated by combing SNP P-values into a gene test-statistic, indicating the association between the gene and the GWAS phenotype. Genes significantly associated with CT at each ROI were identified as exceeding the FDR corrected threshold.

Random Effects in LMM

We tested significance of random effects in LMM using R function lmerTest: rand. Supplemental Table S21 showed random effects of the family nested within acquisition site across 68 regions of interests (ROI). As to common, externalizing-specific and internalizing-specific regions, almost all random effects were significant. It suggested that sites and families do have significant effects on cortical thickness (CT), which should be taken into account.

Case-control analysis encompassing comorbid cases

When we examined CT alterations in three diagnostic families without excluding comorbid cases, CT was significantly higher in externalizing (6 regions, Figure S5A), internalizing (1 region, Figure S5B) and thought disorders (1 region, Figure S5C) compared with healthy controls (HC). Both externalizing and internalizing disorders exhibited thicker CT in left pars opercularis, which is consistent with our result in case-control analysis excluding comorbid cases. Therefore, CT in left pars opercularis was indeed a transdiagnostic biomarker shared by externalizing and internalizing disorders. Other significantly altered regions in externalizing disorders included left posterior cingulate cortex (PCC), bilateral superior temporal gyrus, left pars orbitalis and right middle temporal gyrus, two of which (left superior temporal gyrus and left pars orbitalis) were also identical with the result in case-control analysis excluding comorbid cases. For thought disorders, CT in left rostral anterior cingulate cortex (ACC) was significantly thicker. In summary, by contrast with results in case-control analysis excluding comorbid cases, results here suggested that excluding comorbid cases helped us eliminate confounding effects introduced by comorbidities, which may otherwise cover the distinct CT alterations of diagnostic families.

Association analysis restricted in patients groups

To examine whether patient groups exhibited distinct associations, we also performed association analysis between CT of regions significantly altered in externalizing and internalizing disorders, and CBCL and cognition scores (baseline and 2-year follow-up) restricted to externalizing and internalizing disorders group, separately. False Discovery Rate (FDR) were used for multiple comparisons. For baseline CBCL scores, youth with externalizing disorders showed positive relationships between CT in left superior temporal gyrus and CBCL Aggressive Behavior (p=7.2×10^-4^, t=3.395) and Externalizing Problems (p=0.004, t=2.923). For baseline and 2-year follow-up cognition scores as well as 2-year follow-up CBCL scores, no associations survived FDR correction.

**Fig. S1. Flowchart depicting inclusion/exclusion criteria in case-control analyses at baseline**

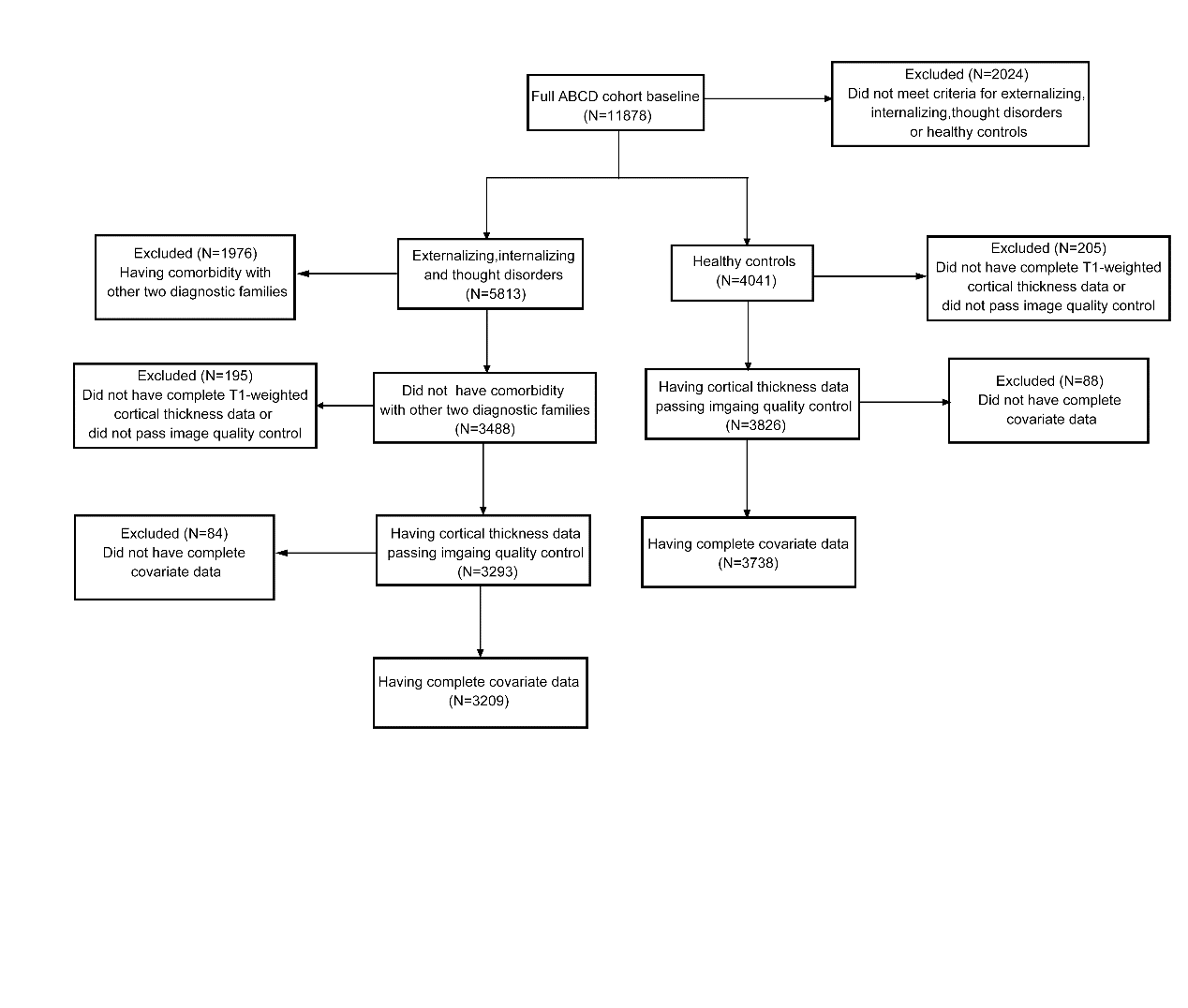

**Fig. S2 Flowchart depicting inclusion/exclusion criteria in Genome-wide association analysis**

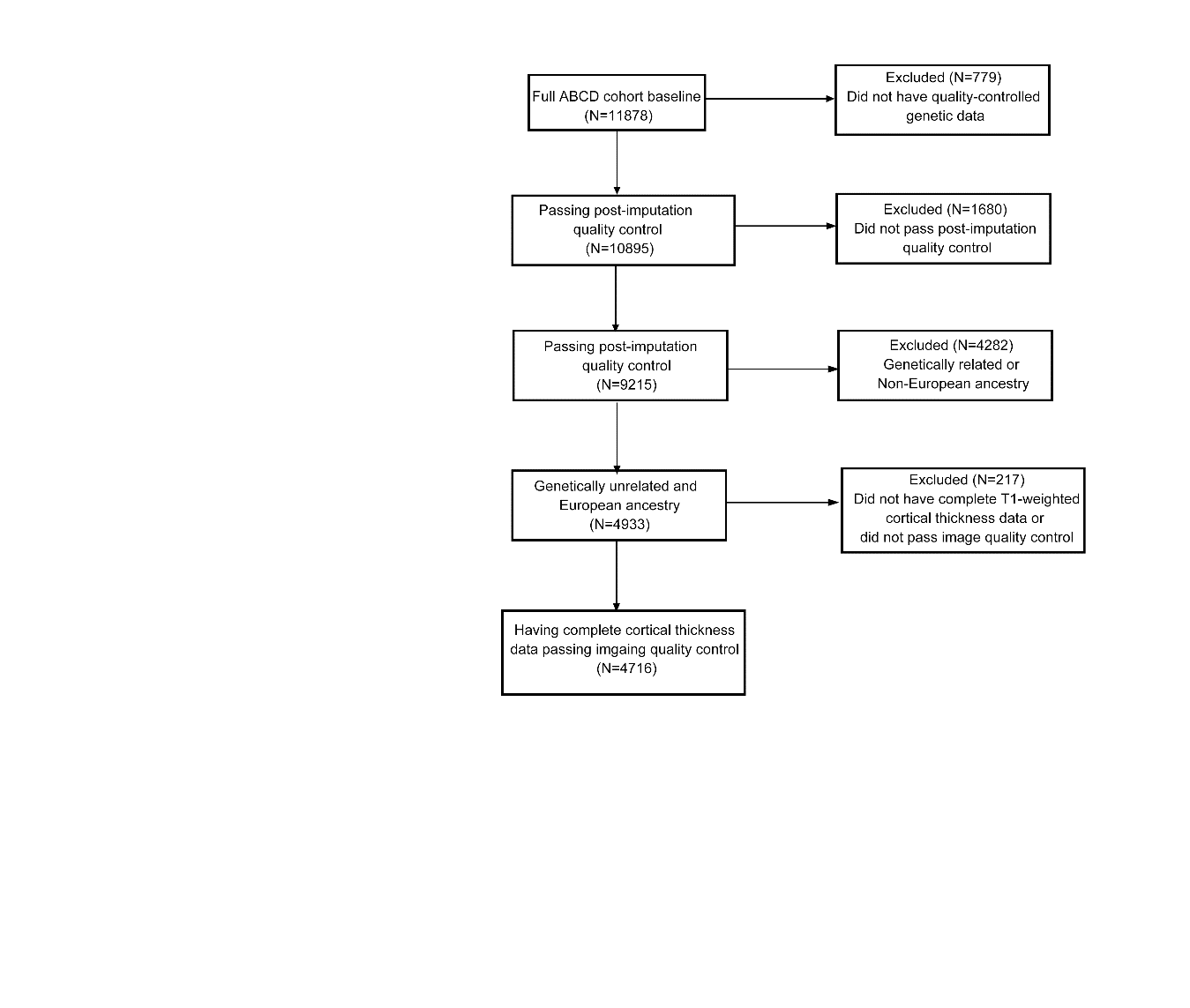

**Fig. S3 Flowchart depicting inclusion/exclusion criteria in association analyses between baseline CT in significantly altered regions and baseline CBCL and cognition scores**

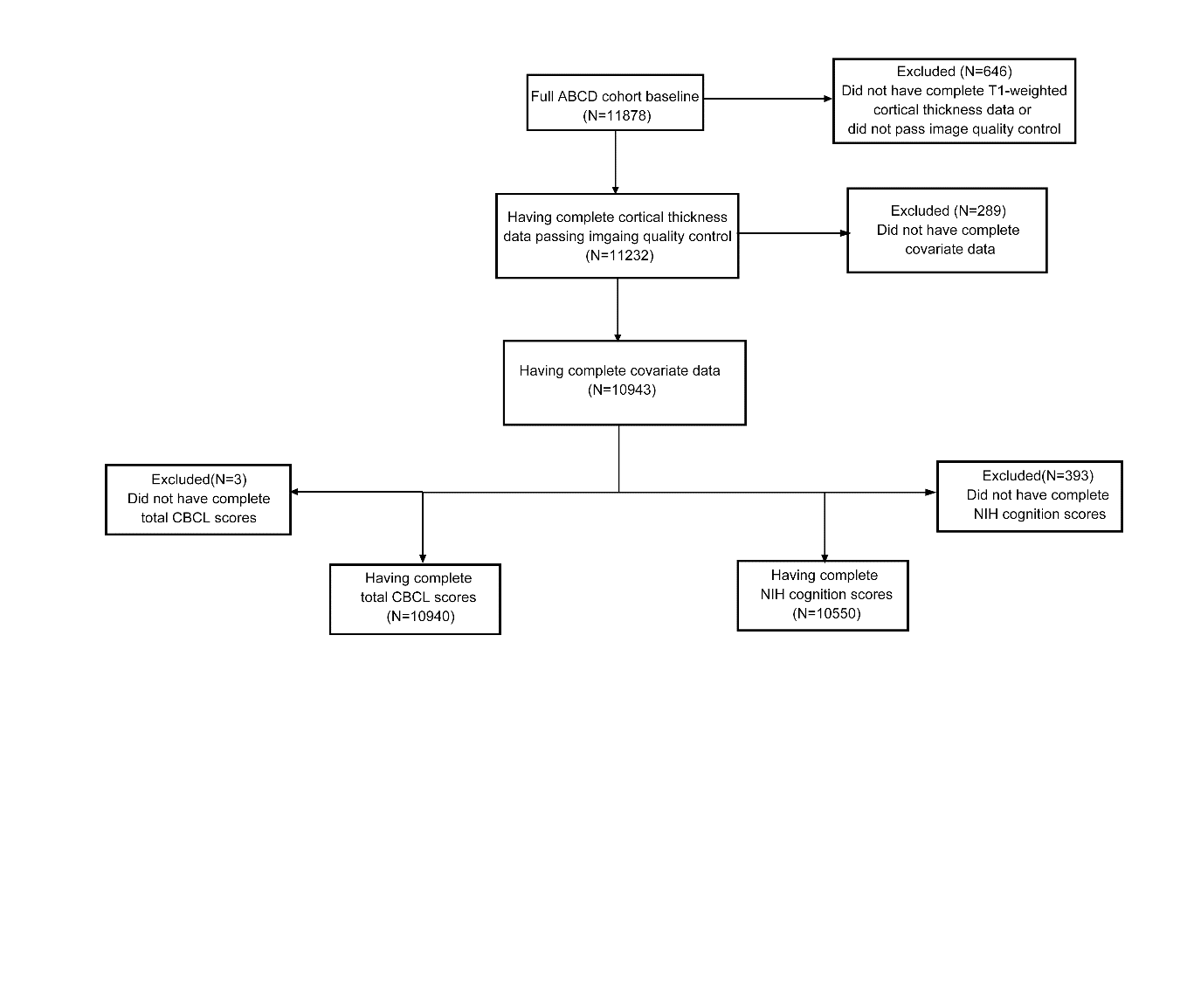

**Fig. S4 Flowchart depicting inclusion/exclusion criteria in association analysis between baseline CT in significantly altered regions and 2-year follow-up CBCL and cognition scores**

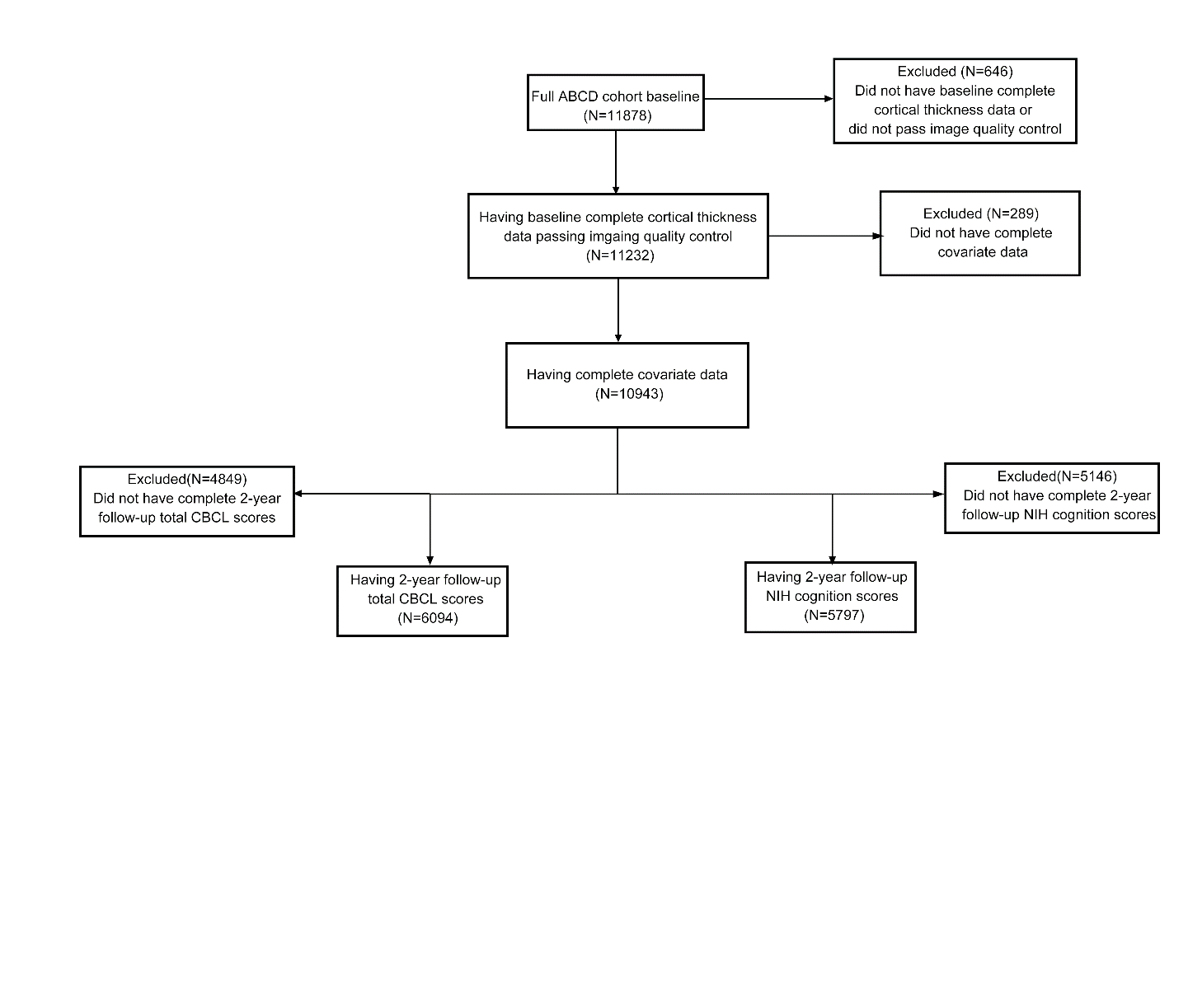

**Fig. S5 Flowchart depicting inclusion/exclusion criteria in longitudinal analysis**

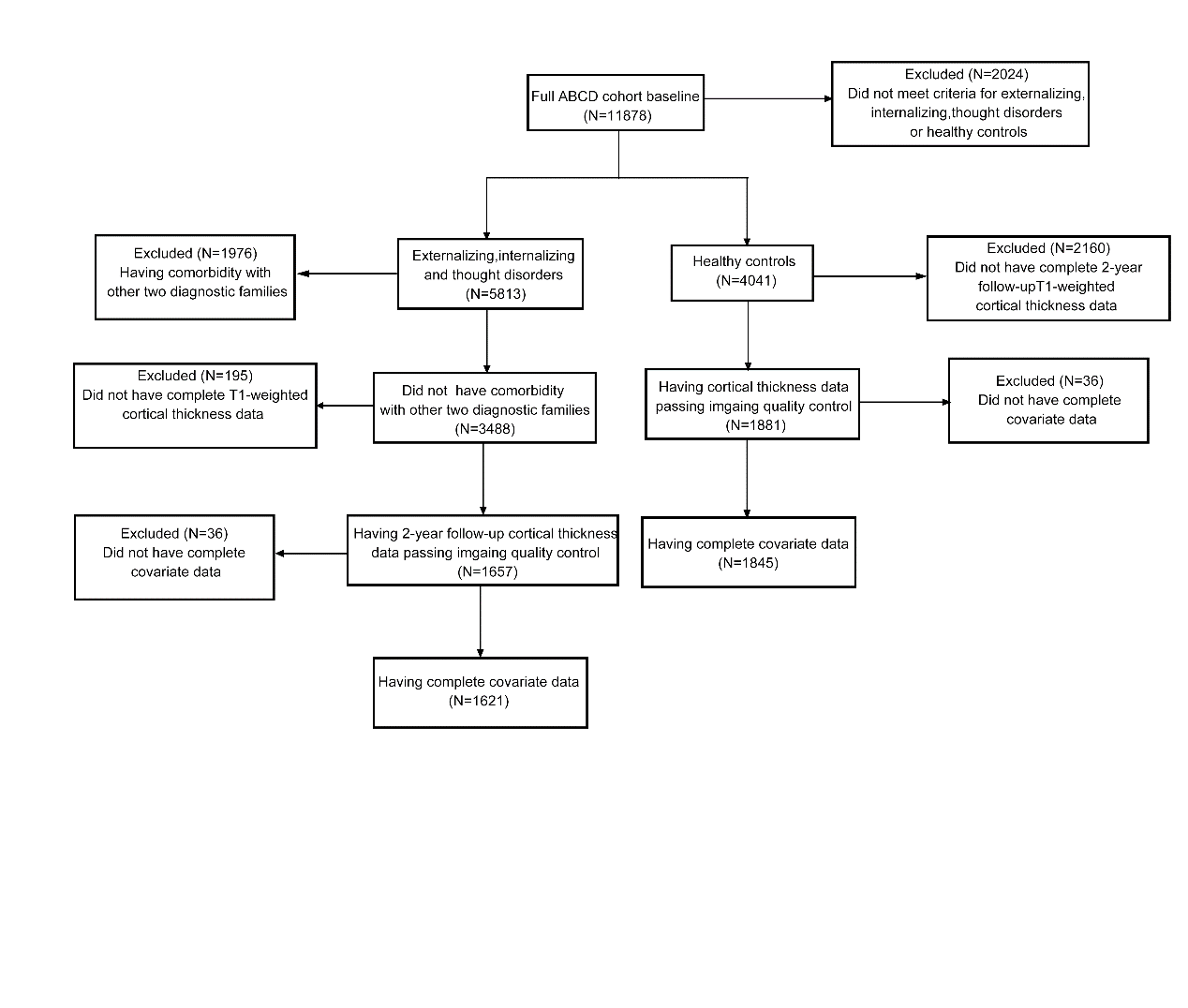

**Fig. S6 Regions showing significant (P_FDR_<0.05) thickness alterations in externalizing, internalizing and thought disorders e****ncompassing comorbid cases at baseline.**

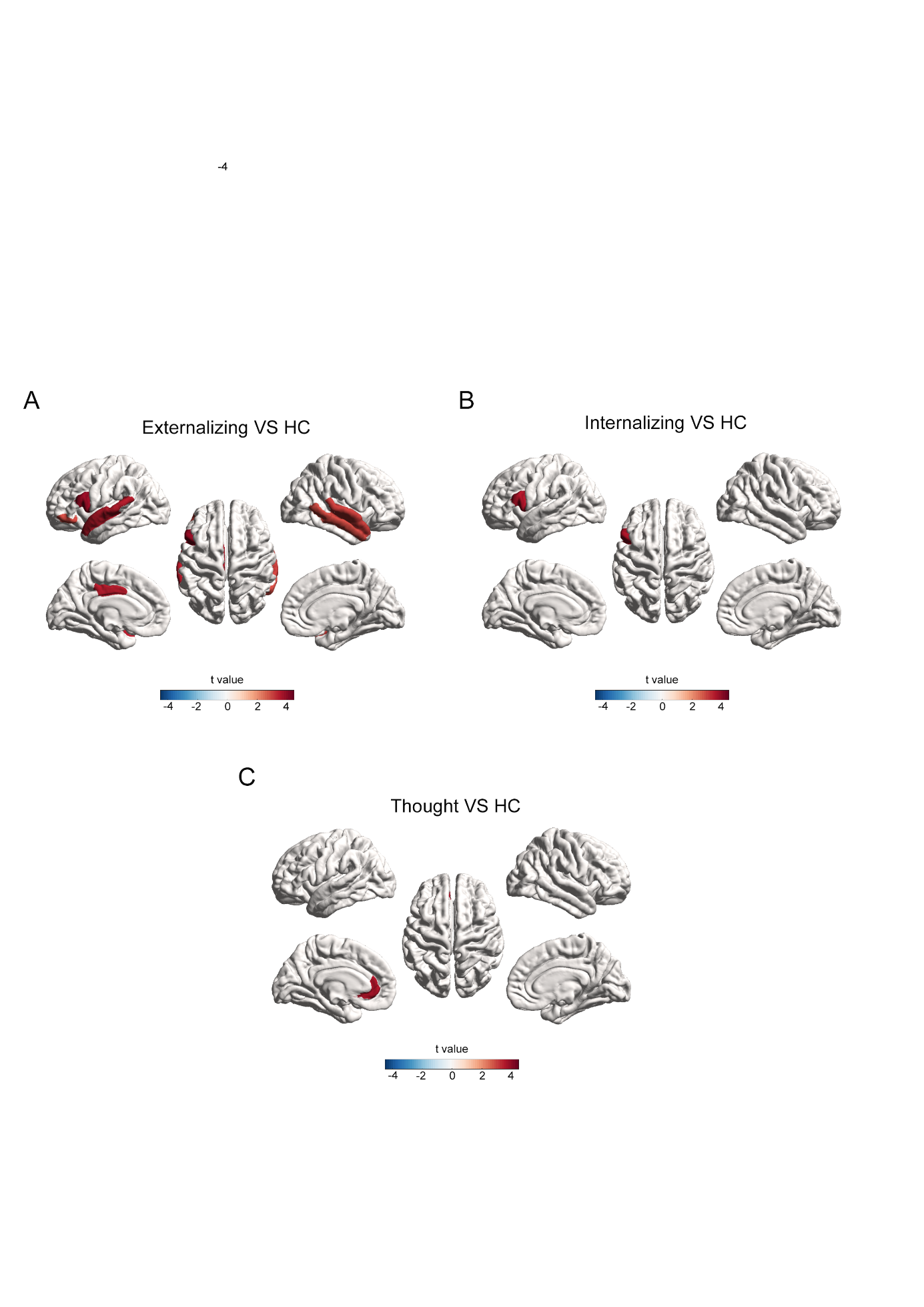

Regions showing significant difference in baseline CT among **A** externalizing, **B** internalizing and **C** thought disorders encompassing comorbid cases, compared with HC group. The colorbars in **A**, **B** and **C** represent the t value of the regression coefficient of group variable from LMM. Abbreviations: Externalizing=externalizing disorders; Internalizing=internalizing disorders; Thought=thought disorders; HC=healthy control.

**Fig. S7 Associations between baseline CT in the Common (CO), Externalizing-specific and Internalizing-specific regions, 2-year follow-up CBCL scores, and 2-year follow-up cognition scores**

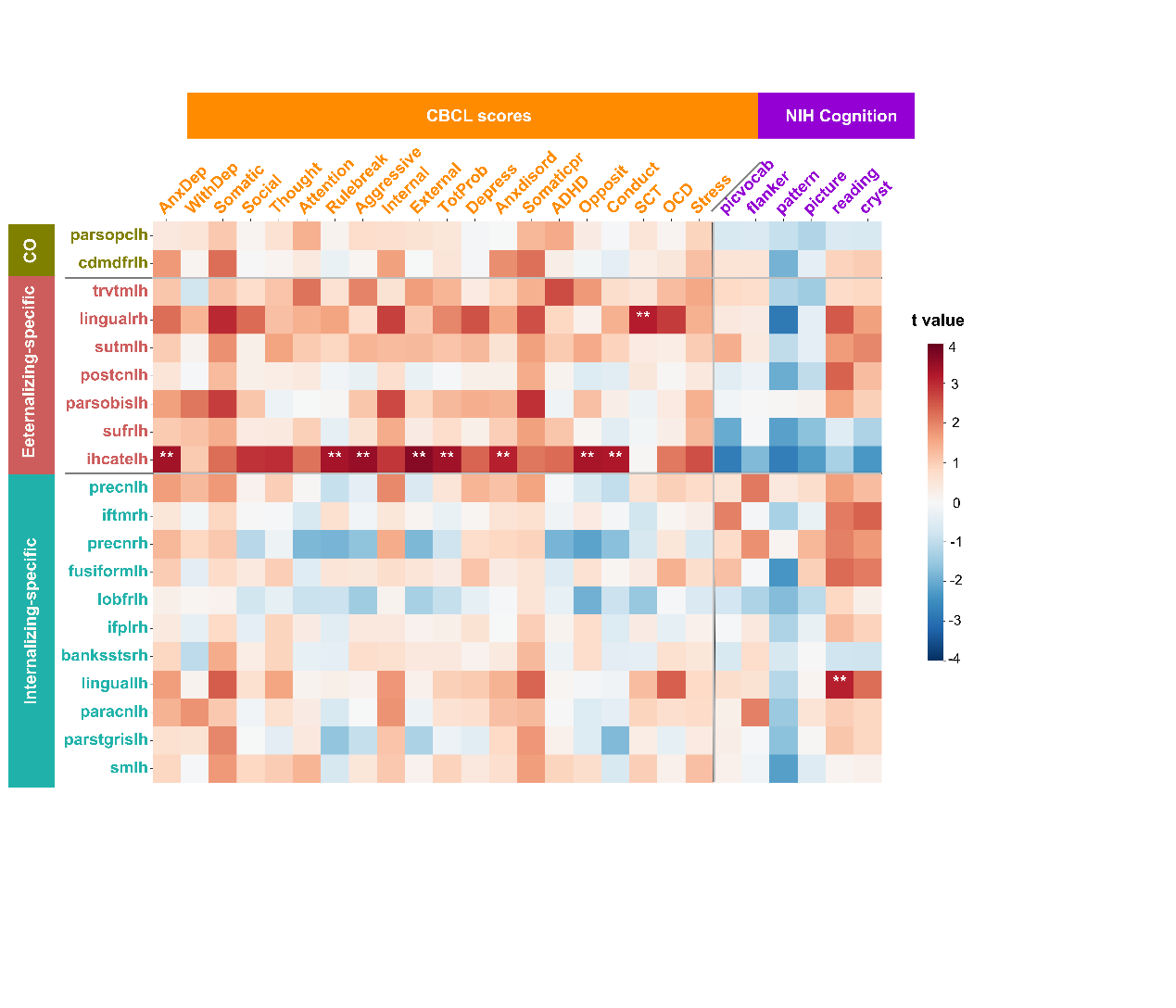

The colorbar represents the t value of the regression coefficient from LMM. Two asterisks (**) indicate P_FDR_<0.05. Abbreviations: parsopclh=left pars opercularis, cdmdfrlh=left caudal middle frontal, trvtmlh=left transverse temporal, lingualrh=right lingual, sutmlh=left superior temporal, postcnlh=left postcentral, parsobislh=left pars orbitalis, sufrlh=left superior frontal, ihcatelh=left isthmus of cingulate cortex, precnlh=left precentral, iftmrh=right inferior temporal, precnrh=right prencentral, fusiformlh=left fusiform, lobfrlh=left lateral orbitofrontal, ifplrh=right inferior parietal lobule, banksstsrh=right banks of superior temporal sulcus, linguallh=left lingual, paracnlh=left paracentral, parstgrislh=left pars triangularis, smlh=left supramarginal, AnxDep=Anxious/Depressed, WithDep=Withdrawn/Depressed, Somatic=Somatic Complaints, Social=Social Problems, Thought=Thought Problems, Attention=Attention Problems, Rulebreak= Rule-Breaking Behavior, Aggressive=Aggressive Behavior, Internal=Internalizing Problems, External=Externalizing Problems, TotProb=Total Problems, Anxdisord=Anxiety disorders, Somaticpr=Somatic Problems, SCT= Sluggish Cognitive Tempo, Opposite= Oppositional Defiant Problems, Conduct=Conduct Problems, OCD=Obsessive-Compulsive Problems, Stress=Stress Problems, ADHD=Attention Deficit/Hyperactivity Problems, picvocab=Picture Vocabulary, flanker=Flanker Inhibitory Control and Attention, list= List Sorting Working Memory, cardsort= Dimensional Change Card Sort, pattern=Pattern Comparison Processing Speed, Picture=Picture Sequence Memory, reading=Oral Reading Recognition, fluidcomp=fluid composite, cryst= crystallized composite, totalcomp=total composite.

**Fig. S8** **Longitudinal changes of CT across the brain from 10 to 12 years old in externalizing, internalizing and thought disorders** **and HC.**

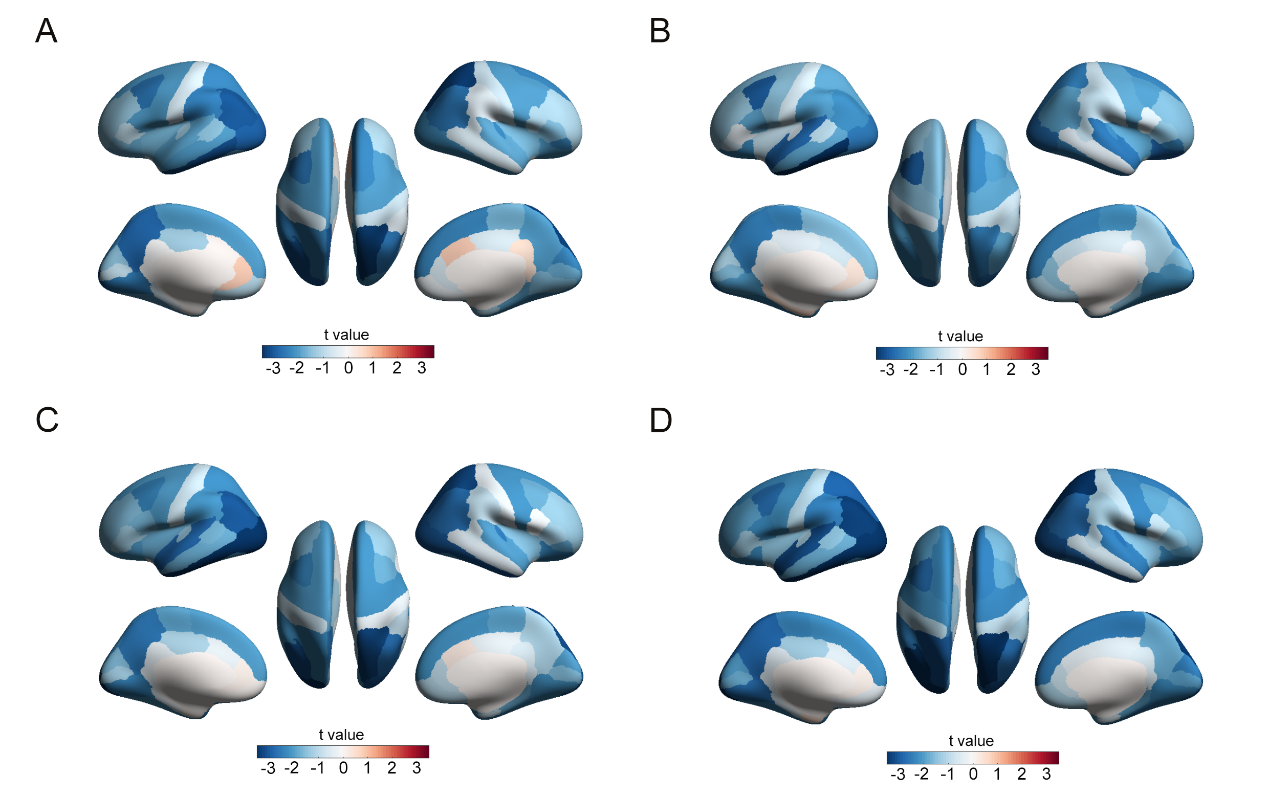

The colorbars in **A**, **B**, **C** and **D** represent t value of the regression coefficient of time variable from LMM.
